# Mendelian randomization analysis of smoking behavior and cognitive ability on the Big Five

**DOI:** 10.1101/2019.12.11.19014530

**Authors:** Charleen D. Adams

## Abstract

Tobacco smoke, a mutagen that can thin the brain’s cortex, might influence the Big Five (neuroticism, conscientiousness, agreeableness, extraversion, and openness). Cognitive ability, however, is a potential confounder, since it is associated with who smokes and with personality. Mendelian randomization (MR), which uses genetic variants as instrumental variables, can be used to probe the causal nature of these factors on personality. Here, MR was used to appraise smoking and cognitive ability on the Big Five and cognitive ability and neuroticism on social disparity. The results seem to suggest that smoking, independent of cognitive ability, leads people to be more neurotic and less extraverted and conscientious. Higher cognitive ability appears to make people less neurotic and more open, when accounting for smoking. Neuroticism appears to increase disparity, and higher cognitive ability appears to decrease it. Smoking may enhance disparity between those of lower and higher cognitive ability by exacerbating personality differences.

---

Personality is a set of enduring traits that influence how individuals interact with their internal, physical, and social environments. Though relatively stable across the lifespan^1^ and highly heritable (heritability estimates range from 33-65%^2–6^), some environmental factors also appear to affect personality^7^. Tobacco smoke may be one of these.

Tobacco smoke has catastrophic effects on human health, including detrimental impacts to tissues outside the respiratory tract^8,9^. For example, tobacco smoke has been documented to thin the brain’s cortex^10^. Further to this, a recent longitudinal study, which adjusted for education (a proxy for cognitive ability), in approximately 15,500 participants found that smoking makes people more neurotic and less agreeable, conscientious, extroverted, and open^11^. However, despite the strong longitudinal design and the adjustment for education, observational studies can still suffer from confounding and reverse causation.

This means that, in this scenario, cognitive ability, a potent predictor of important health and economic outcomes^12,13^, remains a potential confounder (Fig. 1). This is so because a) smoking is done disproportionately by those of lower socioeconomic (SES) status^14^, b) lower SES is a risk factor for lower cognitive ability^15^, c) cognitive ability is associated with various aspects of personality^16,17,26,18–25^, and d) adjustment by cognitive ability in observational studies does not necessarily rule out confounding.

**Fig. 1.**
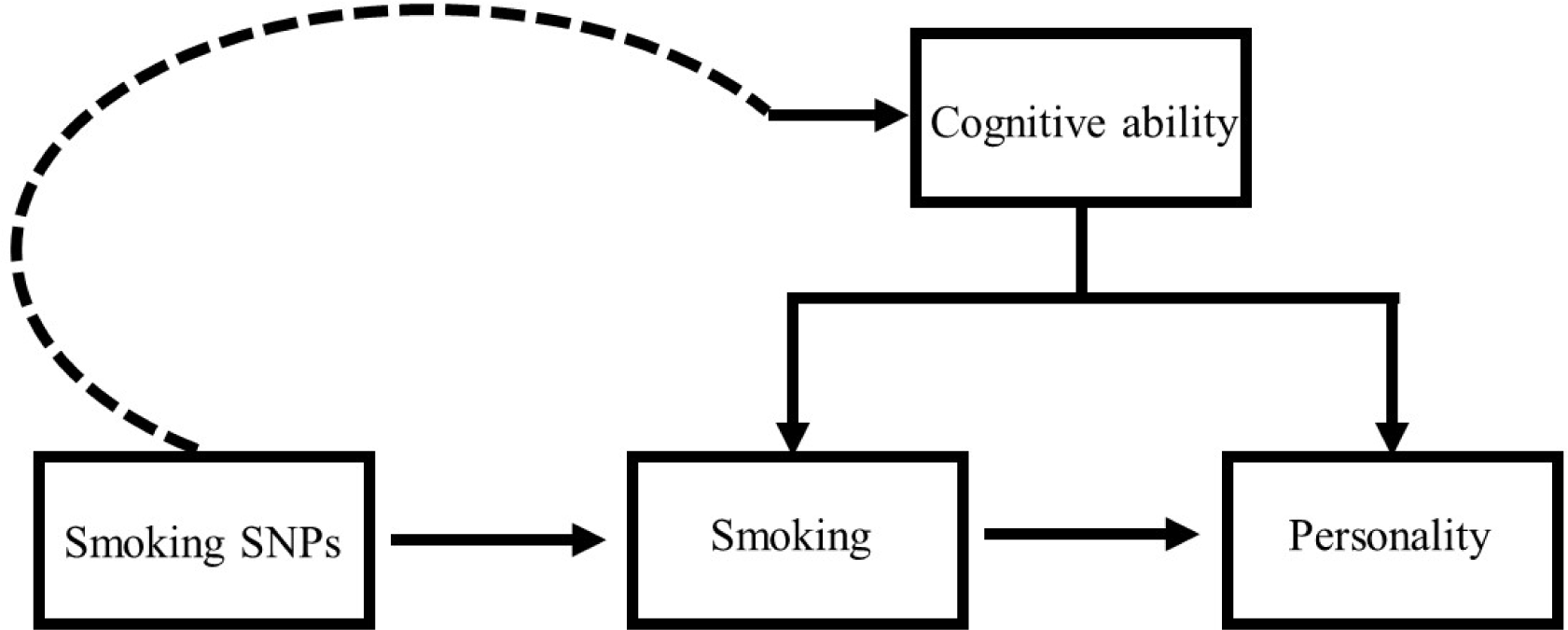
Diagram showing how cognitive ability might explain associations between smoking and personality. This also illustrates possible violations to MR assumptions (ii) and (iii). If the smoking-associated SNPs are also associated with cognitive ability, regardless of whether cognitive ability is a confounder of the relationship between smoking and personality, this would represent a violation to MR assumption (iii); i.e., it would represent a pathway from the smoking SNPs to personality that is independent of smoking. Since cognitive ability is a known potential confounder, if the smoking SNPs used as instrumental variables for smoking are also SNPs for cognitive ability, this is a violation of MR assumption (ii).

Given the known role of cognitive ability on life outcomes and the interwoven relationship between cognitive ability and personality, if smoking causally impacts personality, it may exacerbate disparity between those of different cognitive abilities. Mendelian randomization (MR), a method designed to address confounding and reverse causation, can be used to tease out the causal nature of these relationships, if certain assumptions are met.

## Mendelian randomization

MR is analogous to a randomized-controlled trial (RCT). In an RCT, randomization happens on treatment. With MR, randomization happens on genotype, an exploitation of the random assortment of alleles from parent to offspring. Genetic variants (usually single-nucleotide polymorphisms, SNPs) strongly associated with variables of interest are used as proxies to test the causal impact of environmental exposures: i.e. MR is an instrumental variables technique that uses genetics to understand the environment. Doing so avoids most environmental sources of confounding and forestalls reverse causation, in most circumstances.

MR has three key assumptions, which must hold up in order for the results to be valid: (i) SNPs acting as the instrumental variables must strongly associate with the exposure of interest; (ii) SNPs acting as instrumental variables must be independent of confounders of the exposure and the outcome; and (iii) the SNPs acting as instrumental variables must associate with the outcome of interest only through the exposure (sometimes called the “exclusion restriction”).

## Types of Mendelian randomization

Originally, MR was developed to use summary statistics for genetic variants extracted from a single genome-wide association (GWA) study, but as more GWA studies have been performed and on larger populations, the method was adapted. Two-sample MR adapts the procedure to use summary statistics from two GWA studies^27^. Multivariable MR is a further adaption that enables the inclusion of more than one instrumental variable in a model. This permits adjustment. When univariable and multivariable models are both run, the total (univariable, unadjusted) and direct (multivariable, adjusted) effects can be assessed, as a way to appraise the underlying relationships. Bidirectional MR, as the name suggests, assesses causality in two directions: it is a formal way to test reverse causation. Here, two-sample univariable, multivariable, and bidirectional MR are employed to probe the relationships between smoking, cognitive ability, personality, and disparity.

## Results

### Univariable models of smoking on the Big Five and on cognitive ability (Table 1)

Smoking was associated with an increase in neuroticism (SSGAC): IVW estimate = 0.10; 95% CI = 0.04, 0.16; *P* = 0.002; FDR = 0.007). Likewise, smoking was associated with an increase in neuroticism (GPC): IVW estimate = 0.46; 95% CI = 0.20, 0.71; *P* = 0.004; FDR = 0.002). The sensitivity estimators were similar in direction and magnitude of effects for neuroticism in both the SSGAC and the GPC.

**Table 1.** Causal estimates for smoking on the Big Five and for smoking on cognitive ability (excludes SNPs associated with cognitive ability).

| *Method* | *SNPS* | *B* | *Lower*<br>*95% CI* | *Upper*<br>*95% CI* | *P* | *FDR* | *Q* | *Q*<br>*value* |
| --- | --- | --- | --- | --- | --- | --- | --- | --- |
| **Smoking on neuroticism (SSGAC): n=170,911** |  |  |  |  |  |  |  |  |
| IVW | 65 | 0.10 | 0.04 | 0.16 | 0.002 | 0.007 | 59 | 0.66 |
| MR Egger* | 65 | 0.04 | -0.18 | 0.25 | 0.73 |  | 59 | 0.64 |
| Weighted median* | 65 | 0.07 | -0.02 | 0.17 | 0.14 |  | NA | NA |
| Weighted mode* | 65 | 0.04 | -0.12 | 0.21 | 0.63 |  | NA | NA |
| **Smoking on neuroticism (GPC): n=17,375** |  |  |  |  |  |  |  |  |
| IVW | 47 | 0.46 | 0.20 | 0.71 | 0.0004 | 0.002 | 35 | 0.88 |
| MR Egger* | 47 | 0.77 | -0.14 | 1.69 | 0.10 |  | 35 | 0.87 |
| Weighted median* | 47 | 0.47 | 0.12 | 0.83 | 0.01 |  | NA | NA |
| Weighted mode* | 47 | 0.63 | -0.03 | 1.28 | 0.07 |  | NA | NA |
| **Smoking on conscientiousness (GPC): n=17,375** |  |  |  |  |  |  |  |  |
| IVW | 49 | -0.32 | -0.56 | -0.07 | 0.01 | 0.02 | 44 | 0.62 |
| MR Egger* | 49 | -1.13 | -2.03 | -0.23 | 0.02 |  | 41 | 0.71 |
| Weighted median* | 49 | -0.49 | -0.86 | -0.12 | 0.01 |  | NA | NA |
| Weighted mode* | 49 | -0.52 | -1.07 | 0.04 | 0.07 |  | NA | NA |
| **Smoking on agreeableness (GPC): n=17,375** |  |  |  |  |  |  |  |  |
| IVW | 50 | -0.23 | -0.47 | 0.003 | 0.05 | 0.09 | 37 | 0.90 |
| MR Egger* | 50 | -0.57 | -1.44 | 0.28 | 0.20 |  | 36 | 0.90 |
| Weighted median* | 50 | -0.19 | -0.53 | 0.15 | 0.29 |  | NA | NA |
| Weighted mode* | 50 | -0.27 | -0.83 | 0.29 | 0.36 |  | NA | NA |

| Smoking on extraversion (GPC): n=17,375 |  |  |  |  |  |  |  |  |
| --- | --- | --- | --- | --- | --- | --- | --- | --- |
| IVW | 51 | -0.22 | -0.46 | 0.02 | 0.07 | 0.11 | 44 | 0.71 |
| MR Egger* | 51 | 0.02 | -0.87 | 0.91 | 0.97 |  | 44 | 0.68 |
| Weighted median* | 51 | -0.14 | -0.48 | 0.20 | 0.42 |  | NA | NA |
| Weighted mode* | 51 | -0.19 | -0.78 | 0.41 | 0.54 |  | NA | NA |
| Smoking on openness (GPC): n=17,375 |  |  |  |  |  |  |  |  |
| IVW | 50 | -0.02 | -0.26 | 0.22 | 0.89 | 0.92 | 30 | 0.99 |
| MR Egger* | 50 | 0.09 | -0.80 | 0.97 | 0.85 |  | 29 | 0.98 |
| Weighted median* | 50 | -0.05 | -0.38 | 0.28 | 0.75 |  | NA | NA |
| Weighted mode* | 50 | 0.01 | -0.56 | 0.58 | 0.98 |  | NA | NA |
| Smoking on cognitive ability: n=149,051 |  |  |  |  |  |  |  |  |
| IVW | 54 | -0.14 | -0.28 | 0.01 | 0.07 | 0.11 | 51 | 0.54 |
| MR Egger* | 54 | 0.27 | -0.23 | 0.76 | 0.23 |  | 48 | 0.61 |
| Weighted median* | 54 | -0.04 | -0.24 | 0.16 | 0.70 |  | NA | NA |
| Weighted mode* | 54 | 0.08 | -0.31 | 0.47 | 0.70 |  | NA | NA |

\*Denotes a sensitivity test. $\beta$ =beta estimate; CI=confidence interval; $P$ = $p$ -value; FDR=false-discovery rate corrected $p$ -value; IVW=inverse weighted variance; $Q$ = $Q$ -statistic; $Q$ and $Q$ -value pertain to the Cochrane test for heterogeneity ( $Q$ -value $\geq 0.05$ is evidence against heterogeneity); SSGAC=Social Science Genetic Association Consortium; GPC=Genetics of Personality Consortium.

Smoking was associated with a decrease in conscientiousness: IVW estimate = -0.32; 95% CI = - 0.56, -0.07; *P* = 0.01; FDR = 0.02). The sensitivity estimators were similar in direction, but the magnitude of the MR-Egger estimate was greater than the others, which could imply some pleiotropy.

Smoking did not appear to influence agreeableness: IVW estimate = -0.23; 95% CI = -0.47, 0.003; *P* = 0.05; FDR = 0.09). The sensitivity estimators were similar in direction, but the magnitude of the effect estimate for the MR-Egger was greater than the others.

Smoking did not appear to influence extraversion: IVW estimate = -0.22; 95% CI = -0.46, 0.02; *P* = 0.07; FDR = 0.11). The MR-Egger estimate was in the opposite direction.

Smoking did not appear to influence openness: IVW estimate = -0.02; 95% CI = -0.26, 0.22; *P* = 0.89; FDR = 0.92). The MR-Egger and weighted mode estimators were in the opposite direction from that of the IVW estimate.

Smoking did not appear to influence cognitive ability: IVW estimate = -0.14; 95% CI = -0.28, 0.01; *P* = 0.07; FDR = 0.11). The MR-Egger and weighted mode estimators were in the opposite direction from that of the IVW estimate.

### Univariable models of reverse directions (Table 2)

Neuroticism (SSGAC) did not appear to influence smoking: IVW estimate = 0.10; 95% CI = 0.002, 0.20; *P* = 0.05; FDR = 0.09). The sensitivity estimators were similar in direction, but the magnitude of the effect estimate for the MR-Egger was greater than the others.

**Table 2.** Reverse directions: causal estimates for neuroticism on smoking (excluding SNPs for cognitive ability) and cognitive ability on smoking

| *Method* | *SNPS* | $\beta$ | *Lower*<br>*95% CI* | *Upper*<br>*95% CI* | *P* | *FDR* | *Q* | *Q*<br>*value* |
| --- | --- | --- | --- | --- | --- | --- | --- | --- |
| Neuroticism (SSGAC) on lifetime smoking (n=462,690) |  |  |  |  |  |  |  |  |
| IVW | 4 | 0.10 | 0.002 | 0.20 | 0.05 | 0.09 | 3 | 0.37 |
| MR Egger* | 4 | 1.90 | -0.60 | 4.40 | 0.27 |  | 1 | 0.57 |
| Weighted median* | 4 | 0.03 | -0.15 | 0.21 | 0.74 |  | NA | NA |
| Weighted mode* | 4 | 0.09 | -0.04 | 0.21 | 0.18 |  | NA | NA |
| Cognitive ability on lifetime smoking (n=462,690) |  |  |  |  |  |  |  |  |
| IVW | 39 | -0.05 | -0.06 | -0.03 | 5.64E-09 | 8.18E-08 | 53 | 0.05 |
| MR Egger* | 39 | -0.06 | -0.13 | 0.01 | 0.11 |  | 53 | 0.05 |
| Weighted median* | 39 | -0.05 | -0.07 | -0.03 | 1.36E-05 |  | NA | NA |
| Weighted mode* | 39 | -0.09 | -0.15 | -0.03 | 0.01 |  | NA | NA |

\*Denotes a sensitivity test. $\beta$ =beta estimate; CI=confidence interval; $P$ = $p$ -value; FDR=false-discovery rate corrected $p$ -value; IVW=inverse weighted variance. $Q$ = $Q$ -statistic; $Q$ and $Q$ -value pertain to the Cochrane test for heterogeneity ( $Q$ -value $\geq 0.05$ is evidence against heterogeneity).

Cognitive ability was associated with a decreased risk for smoking: IVW estimate = -0.05; 95% CI = -0.06, -0.03; *P* = 5.64E-09; FDR = 8.18E-08). The sensitivity estimators were similar in direction and magnitude of effects.

### Univariable models of cognitive ability on the Big Five (Table 3)

Cognitive ability was associated with a decreased risk for neuroticism (SSGAC): IVW estimate = -0.03; 95% CI = - 0.05, -0.01; *P* = 0.005; FDR = 0.01). The sensitivity estimators were similar in direction with some minor variability in the magnitude of effects.

**Table 3.** Main effect of cognitive ability on the Big Five.

| *Method* | *SNPS* | $\beta$ | *Lower*<br>*95% CI* | *Upper*<br>*95% CI* | *P* | *FDR* | *Q* | *Q*<br>*value* |
| --- | --- | --- | --- | --- | --- | --- | --- | --- |

| **Cognitive ability on neuroticism (SSGAC): n=170,911** |  |  |  |  |  |  |  |  |
| --- | --- | --- | --- | --- | --- | --- | --- | --- |
| IVW | 58 | -0.03 | -0.05 | -0.01 | 0.005 | 0.01 | 62 | 0.30 |
| MR Egger* | 58 | -0.10 | -0.19 | -0.01 | 0.04 |  | 60 | 0.35 |
| Weighted median* | 58 | -0.02 | -0.05 | 0.01 | 0.15 |  | NA | NA |
| Weighted mode* | 58 | -0.002 | -0.07 | 0.06 | 0.96 |  | NA | NA |
| **Cognitive ability on neuroticism (GPC): n=17,375** |  |  |  |  |  |  |  |  |
| IVW | 49 | -0.03 | -0.10 | 0.04 | 0.43 | 0.50 | 41 | 0.75 |
| MR Egger* | 49 | -0.22 | -0.59 | 0.14 | 0.23 |  | 40 | 0.76 |
| Weighted median* | 49 | -0.01 | -0.11 | 0.10 | 0.91 |  | NA | NA |
| Weighted mode* | 49 | 0.09 | -0.13 | 0.31 | 0.43 |  | NA | NA |
| **Cognitive ability on conscientiousness (GPC): n=17,375** |  |  |  |  |  |  |  |  |
| IVW | 50 | -0.04 | -0.11 | 0.03 | 0.30 | 0.38 | 50 | 0.45 |
| MR Egger* | 50 | 0.04 | -0.32 | 0.40 | 0.84 |  | 49 | 0.42 |
| Weighted median* | 50 | 0.01 | -0.09 | 0.11 | 0.86 |  | NA | NA |
| Weighted mode* | 50 | 0.08 | -0.15 | 0.32 | 0.48 |  | NA | NA |
| **Cognitive ability on agreeableness (GPC): n=17,375** |  |  |  |  |  |  |  |  |
| IVW | 48 | -0.002 | -0.07 | 0.07 | 0.96 | 0.96 | 36 | 0.87 |
| MR Egger* | 48 | 0.01 | -0.33 | 0.35 | 0.96 |  | 36 | 0.85 |
| Weighted median* | 48 | 0.04 | -0.06 | 0.13 | 0.43 |  | NA | NA |
| Weighted mode* | 48 | 0.08 | -0.12 | 0.28 | 0.46 |  | NA | NA |
| **Cognitive ability on extraversion (GPC): n=17,375** |  |  |  |  |  |  |  |  |
| IVW | 50 | -0.05 | -0.11 | 0.02 | 0.17 | 0.22 | 35 | 0.93 |
| MR Egger* | 50 | 0.21 | -0.14 | 0.55 | 0.24 |  | 33 | 0.95 |
| Weighted median* | 50 | -0.07 | -0.16 | 0.03 | 0.16 |  | NA | NA |
| Weighted mode* | 50 | -0.09 | -0.29 | 0.12 | 0.41 |  | NA | NA |
| **Cognitive ability on openness (GPC): n=17,375** |  |  |  |  |  |  |  |  |
| IVW | 49 | 0.13 | 0.06 | 0.19 | 0.0003 | 0.002 | 33 | 0.95 |
| MR Egger* | 49 | 0.08 | -0.27 | 0.43 | 0.66 |  | 33 | 0.93 |
| Weighted median* | 49 | 0.12 | 0.03 | 0.22 | 0.0095 |  | NA | NA |
| Weighted mode* | 49 | 0.10 | -0.09 | 0.30 | 0.31 |  | NA | NA |

\*Denotes a sensitivity test. $\beta$ =beta estimate; CI=confidence interval; $P$ = $p$ -value; FDR=false-discovery rate corrected $p$ -value; IVW=inverse weighted variance; $Q$ = $Q$ -statistic; $Q$ and $Q$ -value pertain to the Cochran test for heterogeneity ( $Q$ -value $\geq 0.05$ is evidence against heterogeneity); SSGAC=Social Science Genetic Association Consortium; GPC=Genetics of Personality Consortium.

Cognitive ability was not associated with neuroticism (GPC): IVW estimate = -0.03; 95% CI = - 0.10, 0.04; *P* = 0.43; FDR = 0.50). The sensitivity estimators displayed some variability magnitude and direction of effects.

Cognitive ability was not associated with conscientiousness: IVW estimate = -0.04; 95% CI = - 0.11, 0.03; *P* = 0.30; FDR = 0.38). The sensitivity estimators displayed some variability in the direction of effects.

Cognitive ability was not associated with agreeableness: IVW estimate = -0.002; 95% CI = - 0.07, 0.07; *P* = 0.96; FDR = 0.96). The sensitivity estimators displayed some variability in magnitude and direction of effects.

Cognitive ability was not associated with extraversion: IVW estimate = -0.05; 95% CI = -0.11, 0.02; *P* = 0.17; FDR = 0.24). The sensitivity estimators displayed some variability in magnitude and direction of effects.

Cognitive ability was associated with an increase in openness: IVW estimate = 0.13; 95% CI = 0.06, 0.19; *P* = 0.0003; FDR = 0.002). The sensitivity estimators mostly aligned in magnitude and direction of effects (the MR-Egger estimator’s effect estimate was slightly smaller).

### Multivariable casual estimates for cognitive ability and smoking on the Big Five (Table 4)

Independent and opposing direct effects were observed for both cognitive ability and smoking on risk for neuroticism (SSGAC) with cognitive ability being protective and smoking increasing risk. For neuroticism (GPC), smoking but not cognitive ability had a direct effect. As for neuroticism in the SSGAC, smoking increased risk for neuroticism in the GPC. Smoking did not have direct effects on agreeableness or openness but directly decreased conscientiousness and extraversion. Cognitive ability did not have a direct effect on conscientiousness, agreeableness, or extraversion, but directly increased openness.

**Table 4.** Multivariable casual estimates for smoking and cognitive ability on the Big Five.

| *Exposures* | *SNPs* | *$\beta$* | *Lower*<br>*95% CI* | *Upper*<br>*95% CI* | *P* | *FDR* |
| --- | --- | --- | --- | --- | --- | --- |
| **Neuroticism (SSGAC): n=170,911** |  |  |  |  |  |  |
| Cognitive ability | 75 | -0.04 | -0.07 | -0.01 | 0.002 | 0.007 |
| Smoking | 82 | 0.12 | 0.03 | 0.21 | 0.007 | 0.018 |

| **Neuroticism (GPC): n=17,375** |  |  |  |  |  |  |
| --- | --- | --- | --- | --- | --- | --- |
| Cognitive ability | 51 | -0.03 | -0.10 | 0.03 | 0.33 | 0.4 |
| Smoking | 54 | 0.48 | 0.24 | 0.72 | 0.00008 | 0.0008 |
| **Conscientiousness (GPC): n=17,375** |  |  |  |  |  |  |
| Cognitive ability | 51 | -0.06 | -0.13 | 0.01 | 0.10 | 0.15 |
| Smoking | 54 | -0.34 | -0.59 | -0.09 | 0.01 | 0.02 |
| **Agreeableness (GPC): n=17,375** |  |  |  |  |  |  |
| Cognitive ability | 51 | -0.01 | -0.07 | 0.05 | 0.74 | 0.79 |
| Smoking | 54 | -0.24 | -0.46 | -0.02 | 0.03 | 0.06 |
| **Extraversion (GPC): n=17,375** |  |  |  |  |  |  |
| Cognitive ability | 51 | -0.04 | -0.11 | 0.02 | 0.17 | 0.22 |
| Smoking | 54 | -0.29 | -0.51 | -0.07 | 0.01 | 0.02 |
| **Openness (GPC): n=17,375** |  |  |  |  |  |  |
| Cognitive ability | 51 | 0.10 | 0.04 | 0.17 | 0.0008 | 0.004 |
| Smoking | 54 | -0.08 | -0.29 | 0.14 | 0.48 | 0.54 |

\*Denotes a sensitivity test. $\beta$ =beta estimate; CI=confidence interval; $P$ = $p$ -value; FDR=false-discovery rate corrected $p$ -value; SSGAC=Social Science Genetic Association Consortium; GPC=Genetics of Personality Consortium.

### Cognitive ability and neuroticism on disparity (Table 5)

Higher cognitive ability was associated with a decrease in disparity: IVW estimate = -0.03; 95% CI = -0.04, -0.02; *P* = 5.22E-09; FDR = 8.18E-08). The sensitivity estimators aligned in magnitude and direction of effects.

**Table 5.**
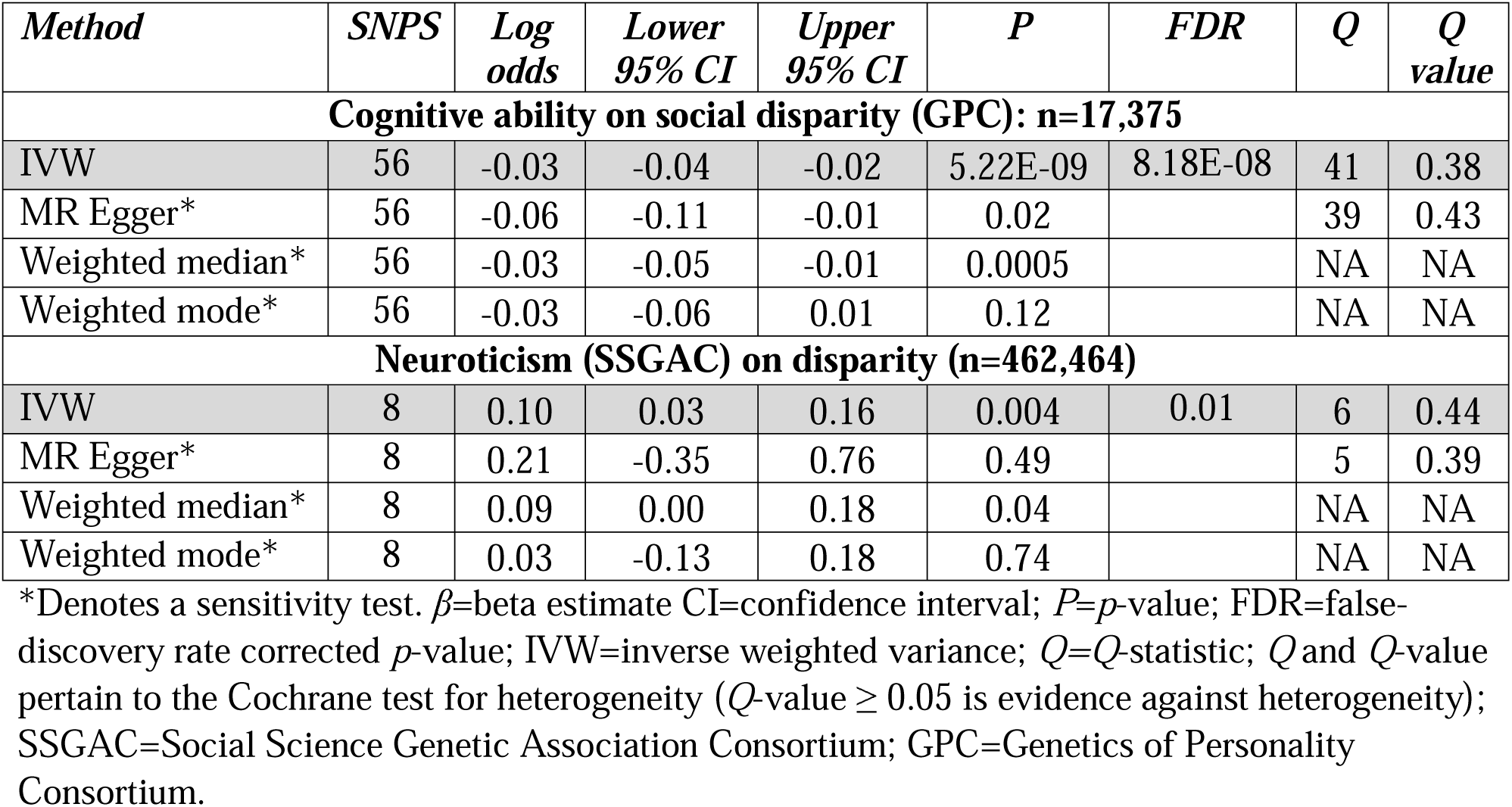
Casual estimates for cognitive ability and neuroticism on social disparity.

Neuroticism (SSGAC) was associated with an increase in disparity: IVW estimate = 0.10; 95% CI = 0.03, 0.16; *P* = 0.004; FDR = 0.01). The sensitivity estimators aligned in magnitude and direction of effects.

Supplementary Tables 2-8, 14-19, 27-33, 40-41, and 44-45 contain the results for individual SNPs.

## Discussion

### Summary of findings

After multiple-testing correction and removing SNPs that influence cognitive ability, in the univariable models of smoking and the Big Five, there was evidence for smoking increasing neuroticism (SSGAC and GPC) and decreasing conscientiousness. After adjusting for cognitive ability, however, there was evidence that smoking directly increases neuroticism and directly decreases conscientiousness and also extraversion. There was suggestive evidence that smoking also decreases agreeableness in the multivariable model prior to, but not after, multiple testing. Smoking did not have an effect on openness or cognitive ability. In both the univariable and multivariable models of cognitive ability on the Big Five, cognitive ability protects against neuroticism (SSGAC) and increases openness. In the univariable models of neuroticism (SSGAC) and cognitive ability on social disparity, neuroticism increases risk for greater social disparity, and higher cognitive ability protects against it.

As hypothesized, higher cognitive ability was observed to decrease smoking behavior. This suggests that cognitive ability is a confounder of the smoking-personality relationships and might, therefore, be a source of horizontal pleiotropy. The tests that show non-zero direct effects of smoking and cognitive ability in the multivariable models, however, suggest that the effects of smoking and cognitive ability estimated by the univariable MR analyses are unlikely to entirely be due to horizontally pleiotropic effects.

Excepting for openness, which appeared not to be affected by smoking, the MR results lend support to the majority of the longitudinal findings by Stephan *et al*. (2019), which found that smoking increases neuroticism and decreases the other Big Five personality traits^11^. Beyond this, the present study suggests that smoking’s influence on personality may enhance social disparity by exacerbating personality differences between those of lower and higher cognitive ability. This further implies that smoking is a potential barrier to economic success and that campaigns and policies for reducing smoking may be beneficial far beyond the common knowledge that not smoking protects from various chronic diseases. This knowledge might be utilized in on-going strategies aimed at reducing smoking.

### Strengths and limitations

The study has a number of limitations. First, MR studies are always subject to the possibility of unwanted pleiotropy, which cannot be entirely ruled out. Measures were taken to investigate this possibility, however, and smoking-associated SNPs related to cognitive ability were removed to reduce the possibility for violations to MR assumptions (ii) and (iii). For the traits showing evidence of causality, substantial bias from pleiotropy was not apparent, though for most of the traits, the MR-Egger test, the most conservative of the sensitivity estimators, did not provide additional evidence to support the IVW.

Second, for the test of cognitive ability on neuroticism, there may be up to 72% overlap in the participants in the GWA studies used in the two-sample MR. This can lead to bias in the estimates that is compounded by “winner’s curse”, an overestimation of the SNP-trait effect in the discovery GWA study^28,29^. For the tests of cognitive ability on social disparity and cognitive ability on smoking, there may be up to 32% overlap in the participants in the GWA studies used in the two-sample MR. For the tests of neuroticism (SSGAC) on smoking and neuroticism on social disparity; however, the possibility for overlap is only at most 23% for both. There should be minimal participant overlap for the two-sample MR tests of cognitive ability on the Big Five (GPC) and smoking on the Big Five (GPC).

The present analysis also has some noteworthy strengths. One is that using SNPs associated with lifetime smoking, which captures smoking status and also cessation, duration, and heaviness (versus a less comprehensive measure), may reduce some possible violations to MR assumption (iii), the exclusion restriction assumption. This is because it is conceivable that a measure of smoking that captures only one or some of these aspects of smoking could violate the exclusion restriction, when a more comprehensive measure of smoking that includes them all does not^30^.

Other strengths of present analysis include the incorporation of multivariable and bidirectional MR to better discern confounding and reverse causation and the capitalizing on the power of large samples to detect effects. The two-sample MR design, in particular, is strong, since, if the findings are biased by weak instruments (“winner’s curse” notwithstanding), the effect estimates would be biased towards the null, reducing concerns for false-positives.

### Implications

Observationally, tobacco smokers are noted to score high on neuroticism and low on conscientiousness^6,31,32^. Supposing the present MR results are not biased towards the direction of potential bias in the observational data, the comportment with the observational studies provides some Bayesian (i.e., prior) support for the present findings for these two traits, in particular. But the present findings extend beyond what has been previously reported. They suggest that the correlation in the observational data between smoking and neuroticism is at least partly due to the effect of smoking and not due primarily to reverse causation, neuroticism leading people to smoke. Moreover, the present findings have implications for society: smoking may change aspects of personality that impact life prospects.

Fortunately, globe over, smoking rates have declined, a trend that is likely to continue^33^. With the foreseeable continuation in smoking decline, there should also be a reduction in smoking-driven changes in personality that may enhance disparity between those of different cognitive abilities.

## Methods

### Data sources for genetic instrumental variables

Genetic instrumental variable for smoking: A publicly available GWA study of lifetime smoking, which adjusted for sex and genotyping chip, containing 462,690 in the UK Biobank was chosen^34^. Lifetime smoking is a novel measure, inclusive of smoking status, smoking duration, heaviness, and cessation: a standard deviation (SD) increase in lifetime smoking is “equivalent to an individual smoking 20 cigarettes a day for 15 years and stopping 17 years ago or an individual smoking 60 cigarettes a day for 13 years and stopping 22 years ago”^34^. From this GWA study, independent (those not in linkage disequilibrium; R^2^ < 0.01) SNPs associated at genome-wide significance (*P* < 5 × 10^−8^) with a SD increase in lifetime smoking were identified.

Genetic instrumental variable for cognitive ability: The UK Biobank appraised fluid intelligence by summing the number of correct answers given to 13 fluid intelligence questions (UK Biobank data-field 20016). Members of the Medical Research Council-Integrative Epidemiology Unit (MRC-IEU) at the University of Bristol used PHESANT to run a GWA study of this fluid intelligence measure (n=149,051)^35^. They treated the variable as an ordered categorical type. Thus, the GWA study results indicate a categorical unit increase in fluid intelligence. The summary data are publicly available on MR-Base (available at http://app.mrbase.org/)^27^.

Independent (those not in linkage disequilibrium; R^2^ < 0.01) SNPs associated at genome-wide significance (*P* < 5 × 10^−8^) with a categorical step increase in cognitive ability were identified.

Genetic instrumental variable neuroticism: The Social Science Genetic Association Consortium (SSGAC) ran a GWA study of neuroticism. For this, summary statistics from 170,911 respondents in the Genetics of Personality Consortium (GPC) (n= 63,661) and UK Biobank were pooled. The neuroticism measure for the UK Biobank participants came from their score on a 12-item version of the Eysenck Personality Inventory Neuroticism scale^36,37^. The summary data are reported in standard deviation (SD) units and available through MR-Base^27^. Independent (those not in linkage disequilibrium; R^2^ < 0.01) SNPs associated at genome-wide significance (*P* < 5 × 10^−8^) with a SD increase in neuroticism were identified.

Detailed characteristics for the genetic instrumental variables are available in Supplemental Tables 8-13, 20-26, 34-39, 42-43, 46-47.

### Data sources—outcome data sets

Outcome data set for neuroticism: The SSGAC GWA study for neuroticism, used to obtain genetic instruments for neuroticism, was also used as the outcome GWA study for the test of cognitive ability on neuroticism. The data are in SD-units and publicly available through MR-Base.

Outcome data set for the Big Five: The GPC ran GWA studies of the Big Five on 17,375 participants of European ancestry, using the Neo Personality Inventory^38^ to measure the traits^6^. The summary data are reported as continuous on arbitrary scales and are available through MR-Base. Standardized betas were calculated by dividing both the betas and the standard errors by the SD of the traits as reported in de Moor *et al*. (2012)^6^.

Outcome data set for smoking: The GWA study of lifetime smoking^34^ (hereafter referred to as “smoking”), used to obtain the genetic instrumental variables for smoking, was also chosen for the outcome data source for the test of cognitive ability on smoking. Standardized betas were calculated by dividing both betas and standard errors by the SD of lifetime smoking in the whole sample (SD=0.694). The full summary data is publicly available at https://data.bris.ac.uk/data/dataset/10i96zb8gm0j81yz0q6ztei23d.

Outcome data set for social disparity: Participants in the UK Biobank were assigned Townsend Deprivation Index scores that were calculated from the national census output areas they lived in immediately prior to enrollment (UK Biobank data-field 189). A GWA study of this Townsend Deprivation Index, a continuous variable, was performed by the MRC-IEU using PHESANT^35^. The variable was first transformed to a normal distribution. The data are reported in SD-units and are publicly available through MR-Base.

### Statistical approach

Changes in the outcome traits were calculated with the inverse-variance weighted (IVW) MR method. The “TwoSampleMR” package^27^ was used to do this. All analyses were performed in R version 3.5.2.

### Assessing possible violations to MR assumptions (iii): horizontal pleiotropy

Sensitivity estimators can be used appraise pleiotropic bias. Three were chosen to complement the primary IVW causal tests: MR Egger regression, weighted median, and weighted mode estimations. When the sensitivity estimators comport with the IVW method’s in terms of the direction and magnitude of their effect estimates, this provides some evidence against pleiotropy. This is so because the different sensitivity estimators make different assumptions about the underlying nature of pleiotropy; it is unlikely for them to align if pleiotropy is introducing bias. Thorough descriptions of the various MR sensitivity estimators and the assumptions they make about pleiotropy are provided elsewhere^39–41^.

In addition, since variability in the causal estimates between SNPs can also indicate unwanted pleiotropy, MR radial regression was used to identify SNP outliers^42^. For all meta-analyzed genetic instruments, only those with no outliers were selected for final analysis and report. An additional test for heterogeneity was performed using Cochrane’s *Q*-statistic^43^ on all final genetic instruments (*P-*values ≥ 0.05 indicate a lack of heterogeneity in the SNP effect estimates).

A differing number of SNPs were used to genetically instrument smoking (and cognitive ability) for the Big Five. This is because, in order for a SNP to be included in the meta-analyzed genetic instrument, it had to be available in the outcome GWA study and not be an outlier.

### Assessing violations to MR assumption (ii: confounding)

In order for a trait to be a confounder, it must be associated with both smoking and personality. Cognitive ability fits this criterion, as it is negatively associated with smoking behavior^16–20^ and with neuroticism^21^; both positively and negatively associated, depending upon the study, with conscientiousness^22–24^; and positively associated with both extroversion and openness (specifically crystalized^44^ rather than fluid intelligence^45^)^25,26^. Aspects of agreeableness, but not the trait itself are associated with cognitive ability (e.g., aggression negatively associates with cognitive ability)^21^.Thus, cognitive ability is a highly plausible candidate for confounding, a violation to MR assumption (ii) above (Fig 1).

In addition, despite the statistical checks-and-balances provided by the statistically based MR sensitivity estimators, cognitive ability could violate MR assumption (iii) if the SNPs that instrument smoking are also associated with cognitive ability. To address this, SNPs proxying for smoking and SNPs proxying for fluid intelligence (as measured in a GWA study of fluid intelligence performed by the Neale lab)^46^ were checked for linkage disequilibrium (LD)— whether any of the SNPs were physically correlated. SNPs in LD were removed. The remaining smoking-associated SNPs were run through PhenoScanner, a curated database of GWA studies containing SNP-phenotype associations^47,48^. This identified all known traits with which the smoking SNPs are associated. SNPs found to be associated with any measure of educational attainment (e.g. education years or school qualifications) or cognitive ability (any formal measure of intelligence) were removed. The MR results for smoking on the Big Five including SNPs also tagging cognitive ability are available in Supplemental Tables 2-7, but the results removing SNPs linked with cognitive ability are presented in Table 1.

To formally appraise the direct effects of both cognitive ability and smoking on the Big Five, multivariable analyses including both cognitive ability and smoking (with cognitive ability SNPs removed) were performed for neuroticism (SSGAC) and each of the Big Five traits in the GPC. The SNPs included in these models were not checked for outliers with Radial MR regression nor were SNPs in LD between cognitive ability and smoking removed, since this could lead to a loss in precision for the effect estimates in the multivariable setting^49^. These results are reported in the Abstract.

### Formally assessing reverse causation

Because germline genotypes are fixed (assigned at conception), they temporally precede most other variables under consideration. Thus, use of SNP proxies generally precludes reverse causation^50^; that is, if the MR results indicate causal associations, the direction tested is usually what is responsible for them. There are exceptions, though. One exception would be the following scenario: Say the SNPs genetically instrumenting smoking are not associated with personality except through smoking, but some aspects of personality are developmental, such that budding personality at one stage in life influences smoking behavior, which in turn impacts “final” personality. This is conceivable.

Since the GWA for neuroticism (SSGAC) contains suitable genetic proxies, a bidirectional appraisal was done to assess whether neuroticism causes smoking. Unfortunately, the GWA studies for the Big Five performed by the GPC are not suitable for bidirectional MR, either due to the associations between the Big Five traits and SNPs being too statistically weak (which would violate MR assumption (i)) or not having more than one strongly associated SNP).

A special criterion for bidirectional MR appraisals is that the SNPs instrumenting each trait are not overlapping or in linkage disequilibrium (LD)^51,52^. Therefore, the SNPs instrumenting both smoking and neuroticism (SSGAC) were checked for overlap and LD. As with the primary investigation examining smoking on the Big Five, cognitive ability is still a lurking, highly likely confounder and source of horizontal pleiotropy under this scenario. As such, the smoking SNPs not in LD with fluid intelligence and not associated with any measure of cognitive ability or educational attainment, as assessed by PhenoScanner, were used to instrument smoking in the bidirectional appraisal: neuroticism on smoking.

In addition, cognitive ability on smoking was assessed (comprising a bidirectional MR test along with the association of smoking on cognitive ability). If cognitive ability is observed to influence smoking, this will strengthen the possibility that cognitive ability is a confounder of the smoking-personality relationships, possibly inducing horizontal pleiotropy^53^. As for the assessment of smoking on the Big Five, smoking-associated SNPs associated with cognitive ability were removed, as were any SNPs in LD between smoking and cognitive ability.

### Impact

One criterion for assessing whether potential smoking-shifted changes in personality are important for health is knowing whether aspects of personality and factors associated with it increase social disparity. As such, the cognitive traits that could be adequately instrumented (neuroticism in the SSGAC and cognitive ability) were examined against the Townsend Deprivation Index, a measure of social disparity.

### Power

Estimates of the proportion of variance in exposures explained by genetic instruments (R^2^), used in the calculation of the *F*-statistic (the strength of the association between genetic instruments and exposure traits) were generated. *F*-statistics <10 are considered to suffer from weak-instrument bias. *F*-statistics are used to assess whether a genetic instrument has power to reject the null of no association. For instance, the instrument for smoking on neuroticism (SSGAC) has an R^2^ = 0.008 and the *F*-statistic = 15. R^2^ and *F*-statistics for all traits are available in Supplemental Tables 14-19, 27-33, 40-45. All genetically instrumented traits had *F*-statistics >10.

In addition, the study was formally powered on neuroticism in the SSGAC (its GWA study contains approximately 100,000 more participants than the GPC GWA studies for the Big Five, though they are also aptly powered). To perform a power calculation for quantitative traits for MR, an educated guess about the true causal effect, the observational association between the traits (for neuroticism = 0.18, obtained from Das *et al*.^54^), the R^2^ (for neuroticism = 0.008), the sample size for the outcome trait, and the variances for the exposure and outcome are required (for neuroticism: both = 1, since both are in standard deviation units). Das *et al*. (2015) displayed the observational estimate for neuroticism as an odds ratio (OR; 1.2). To obtain the beta estimate, the following was done: log(1.2)=0.18 (natural log in R). The mRnd MR power calculator (available at http://cnsgenomics.com/shiny/mRnd/)^55^ was used for the calculation.

### Multiple testing

Counting the MR tests of smoking on neuroticism (SSGAC) and smoking on the Big Five (GPC), smoking on cognitive ability, cognitive ability on neuroticism (SSGAC) and cognitive ability on the Big Five (GPC), reverse tests for neuroticism and cognitive ability on smoking, the multivariable analyses of smoking and cognitive ability on neuroticism (SSGAC) and the Big Five (GPC), and the tests of cognitive ability and neuroticism on disparity, 29 tests were performed. As such, a false-discovery rate (FDR) correction for 29 tests was applied to the IVW estimates.

## Data Availability

All data are publicly available:
http://app.mrbase.org/
https://data.bris.ac.uk/data/dataset/10i96zb8gm0j81yz0q6ztei23d

